# Using deep learning to predict age from liver and pancreas magnetic resonance images allows the identification of genetic and non-genetic factors associated with abdominal aging

**DOI:** 10.1101/2021.06.24.21259492

**Authors:** Alan Le Goallec, Samuel Diai, Sasha Collin, Jean-Baptiste Prost, Théo Vincent, Chirag J. Patel

## Abstract

With age, abdominal organs and tissue undergo important changes. For example, liver volume declines, fatty replacement increases in the pancreas, and patients become more vulnerable to age-related diseases such as non-alcoholic fatty liver disease, alcoholic liver disease, hepatitis, fibrosis, cirrhosis, type two diabetes, cancer, gallstones and inflammatory pancreatic disease. Detecting early abdominal aging and identifying factors associated with this phenotype could help delay the onset of such diseases. In the following, we built the first abdominal age predictor by training convolutional neural networks to predict age from 45,552 liver magnetic resonance images [MRIs] and 36,784 pancreas MRIs (R-Squared=73.3±0.6; root mean squared error=3.70±0.03). Attention maps show that the prediction is driven not only by liver and pancreas anatomical features, but also by surrounding organs and tissue. We defined accelerated abdominal aging as the difference between abdominal age and chronological age, a phenotype which we found to be partially heritable (h_g^2^=26.3±1.9%). Accelerated abdominal aging is associated with seven single nucleotide polymorphisms in six genes (e.g PNPT1, involved in RNA metabolic processes). Similarly, it is associated with biomarkers (e.g body impedance), clinical phenotypes (e.g chest pain), diseases (e.g hypertension), environmental (e.g smoking) and socioeconomic (e.g education) variables, suggesting potential therapeutic and lifestyle interventions to slow abdominal aging. Our predictor could be used to assess the efficacy or emerging rejuvenating therapies on the abdomen.

## Background

With age, the different abdominal organs and tissues undergo important changes ^1^. For example, the liver changes both at the cellular (e.g hepatocyte volume, polyploidy, accumulation of dense bodies, reduced smooth endoplasmic reticulum, reduced number of mitochondria) and the macroscopic (e.g reduced volume by 20-40%, up to 35% reduced blood flow) levels, becoming more vulnerable to age-related liver diseases such as liver fibrosis, non-alcoholic fatty liver disease, alcoholic liver disease, and hepatitis C ^2, 3^. Similarly, the pancreas undergoes fibrosis, atrophies, becomes fattier and vulnerable to age-related pancreas-diseases, leading to age related-pancreas disorders such as diabetes, cancer, gallstones and inflammatory pancreatic disease ^4–6^. Other organs, such as the gastrointestinal tract, undergo similar processes ^7^.

Biological age predictors can help understand the etiology of abdominal aging, with the hope to delay the onset of the aforementioned age-related diseases, and others. Biological age represents the state of the body of an individual and it is the true underlying cause of age-related diseases. It is in contrast with chronological age --commonly referred to as age-- the time since the individual’s birth. Biological age predictors are typically built by training machine learning models to predict chronological age. The prediction outputted by the model can then be interpreted as the individual’s biological age. Predictors have already been built on diverse organ datasets such as brain magnetic resonance images [MRIs] ^8^, heart MRIs ^9^, electrocardiograms ^9, 10^, carotid ultrasound images ^11^, pulse wave analysis records ^11^, full body X-ray images ^12, 13^, chest X-ray images ^14^, eye fundus images ^15^, facial features ^16^, blood samples^17^, DNA methylation ^18^, transcriptomics ^19^, proteomics ^20^, microbiome ^21, 22^ and physical activity measurements ^23^. However, to our knowledge, abdominal MRIs such as liver and pancreas MRIs have never been used to predict age as of this writing.

In the following, we built the first abdominal age predictor. We leveraged 45,552 liver MRIs and 36,784 pancreas MRIs (Figure 1A) collected from UK Biobank ^24^ participants aged 37-82 year-old and trained deep convolutional neural networks to predict age from these datasets. We then performed a genome wide association study [GWAS] to estimate the heritability of accelerated abdominal aging and to identify single nucleotide polymorphisms [SNPs] associated with this phenotype. Similarly, we performed an X-wide association study [XWAS] to identify biomarkers, clinical phenotypes, diseases, environmental and socioeconomic variables associated with accelerated abdominal aging. (Figure 1B)

**Figure 1:**
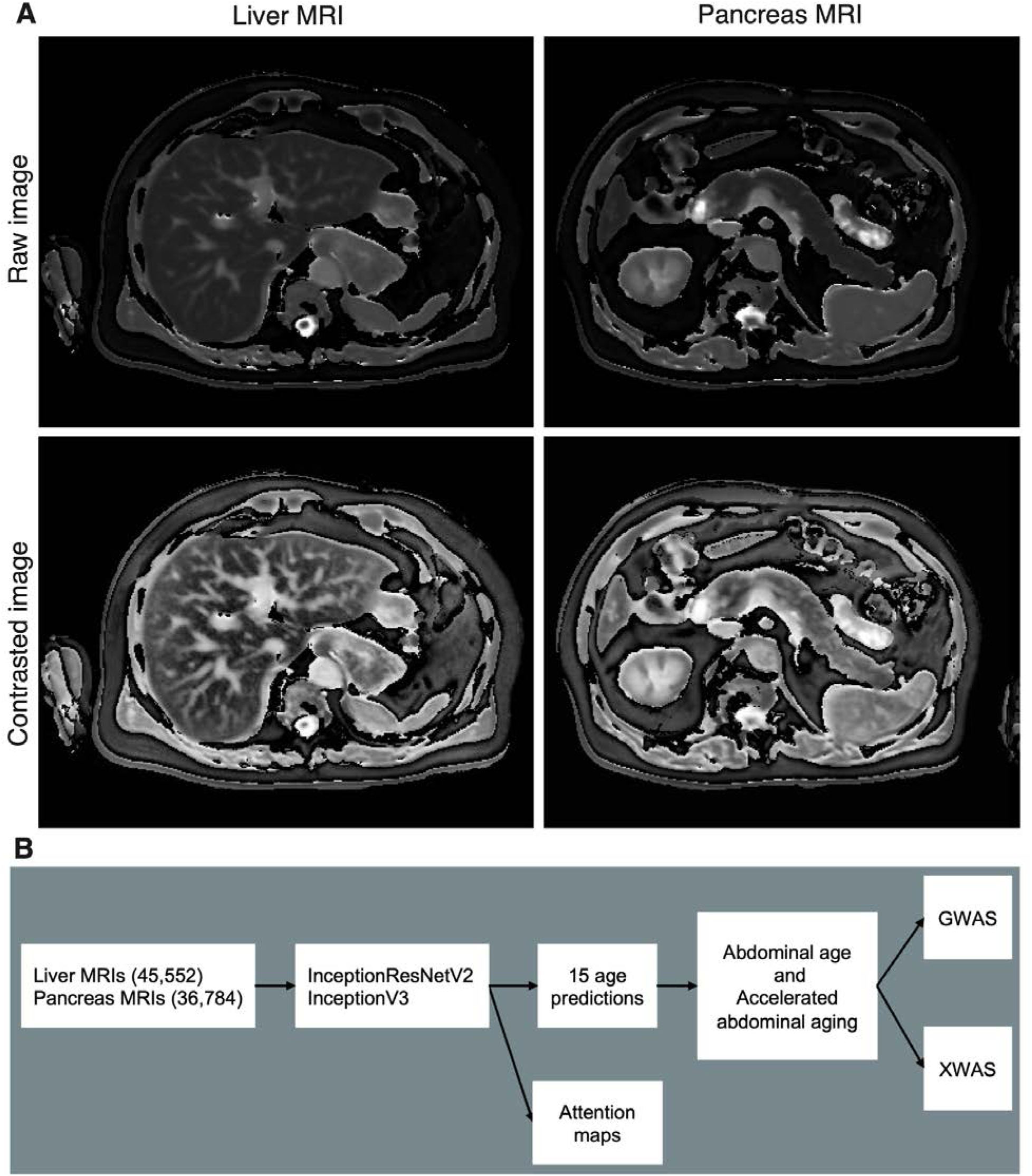
Overview of the datasets and analytic pipeline. A - Sample liver and pancreas MRI images, both raw and preprocessed with a contrasting filter. B - Analytic pipeline.

## Results

### We predicted age within less than four years

We leveraged the UK Biobank, a dataset containing 48,067 liver MRIs and 39,940 pancreas MRIs (Figure 1A) collected from participants aged 37-82 years (Fig. S1). After filtering out low quality images, we used deep convolutional neural networks and transfer learning to predict age from 45,552 liver MRIs (R-Squared [R^2^]=71.5±0.6%; root mean square error [RMSE]=4.09±0.05 years) and from 36,784 pancreas MRIs (R^2^=70.3±0.8; RMSE=4.14±0.04 years), which we then combined into an ensemble model that predicted age with a R^2^ of 76.3±0. and a RMSE of 3.70±0.03 years. (Figure 2)

**Figure 2:**
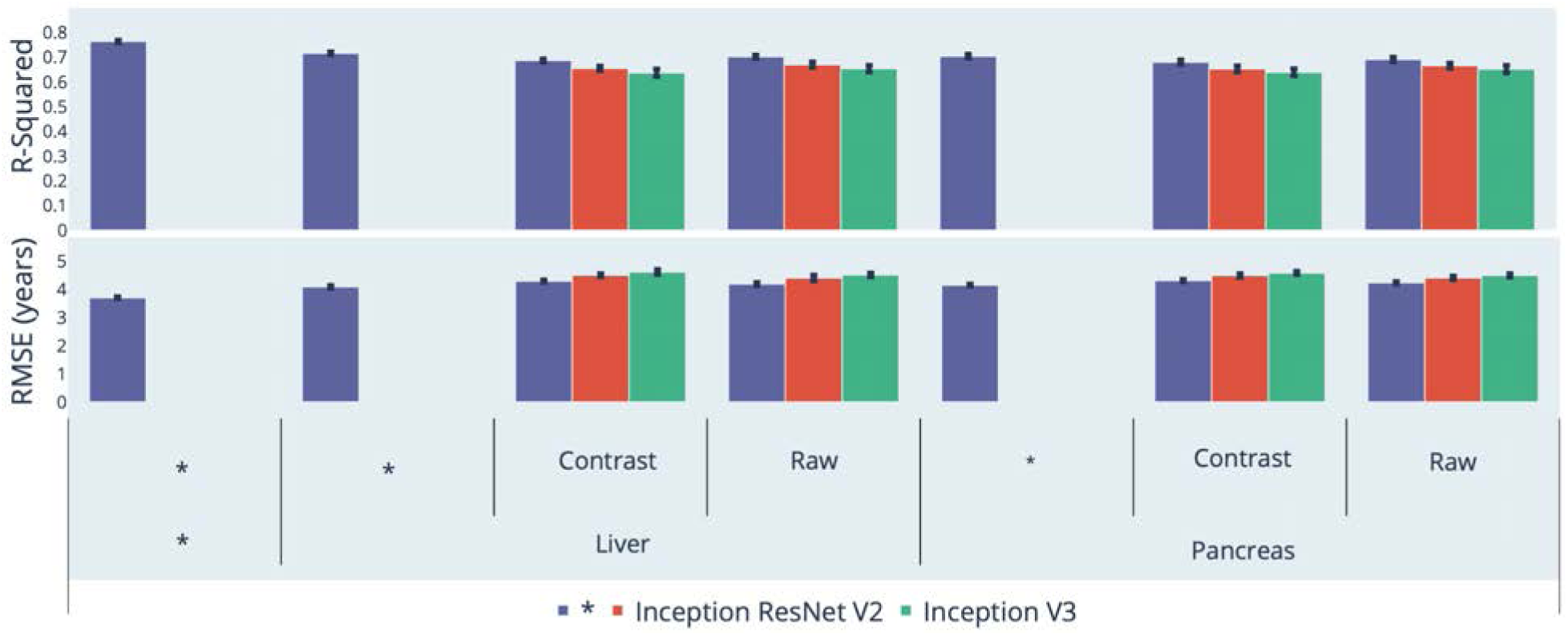
Chronological age prediction performance (R^2^ and RMSE) * represent ensemble models

We defined liver age as the prediction outputted by the liver MRIs-based model, pancreas age as the prediction outputted by the pancreas MRIs-based model, and abdominal age as the prediction outputted by the ensemble model leveraging both liver and pancreas MRIs. All predictions were corrected for the analytical bias in the age prediction residuals (see Methods).

### Identification of features driving abdominal age prediction

For liver MRI-based models, attention maps highlighted the liver along with other abdominal structures such as the stomach, the spleen, muscle and adipose tissue (Figure 3). Similarly, for pancreas MRI-based models, attention maps did not particularly focus on the pancreas but instead highlighted diverse abdominal regions across participants, including the liver (Figure 4).

**Figure 3:**
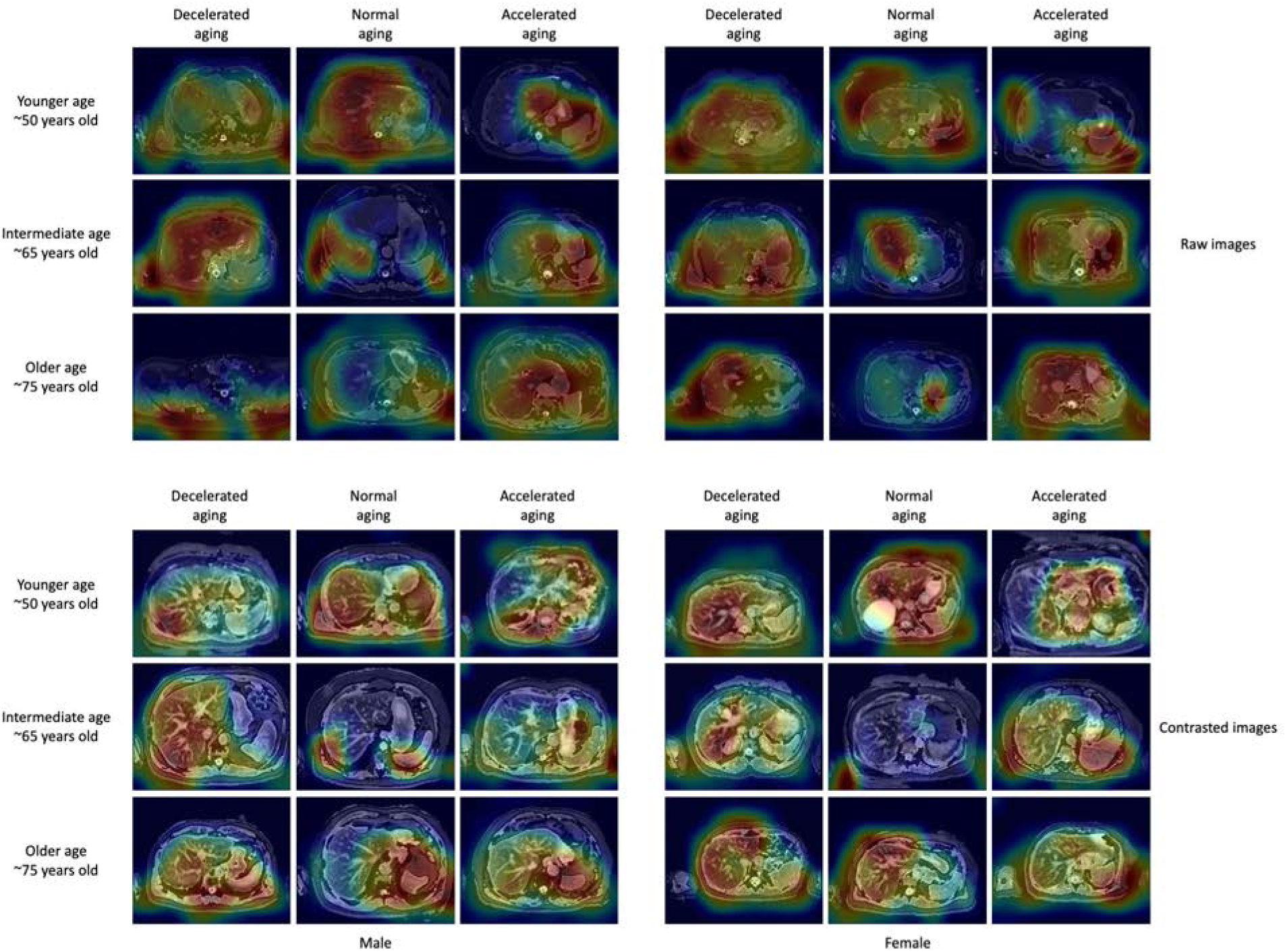
Sample attention maps for liver MRI-based models. Warm filter colors highlight regions of high importance according to the Grad-RAM map.

**Figure 4:**
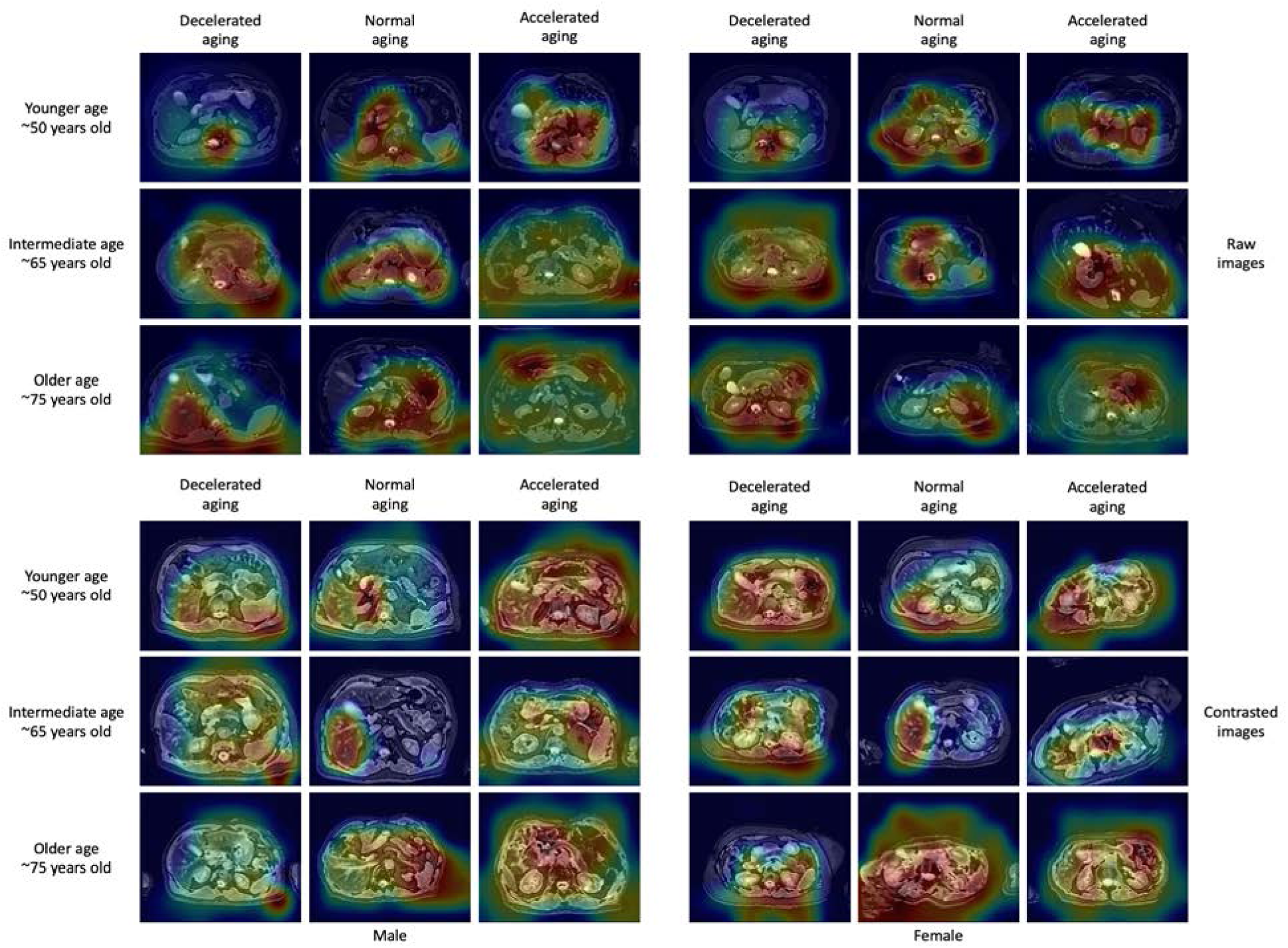
Sample attention maps for pancreas MRI-based models. Warm colors highlight regions of high importance according to the Grad-RAM map.

### Genetic factors and heritability of accelerated abdominal aging

We performed three genome wide association studies [GWASs] to estimate the GWAS-based heritability of abdominal (h_g^2^=26.3±1.9%), liver MRI-based (h_g^2^=22.3±1.5%), and pancreas MRI-based (h_g^2^=22.1±1.9%) accelerated aging. We identified seven SNPs in six genes associated with accelerated aging in at least one abdominal dimension (Figure 5, Table S1, Fig. S2 and Fig. S3).

**Figure 5:**
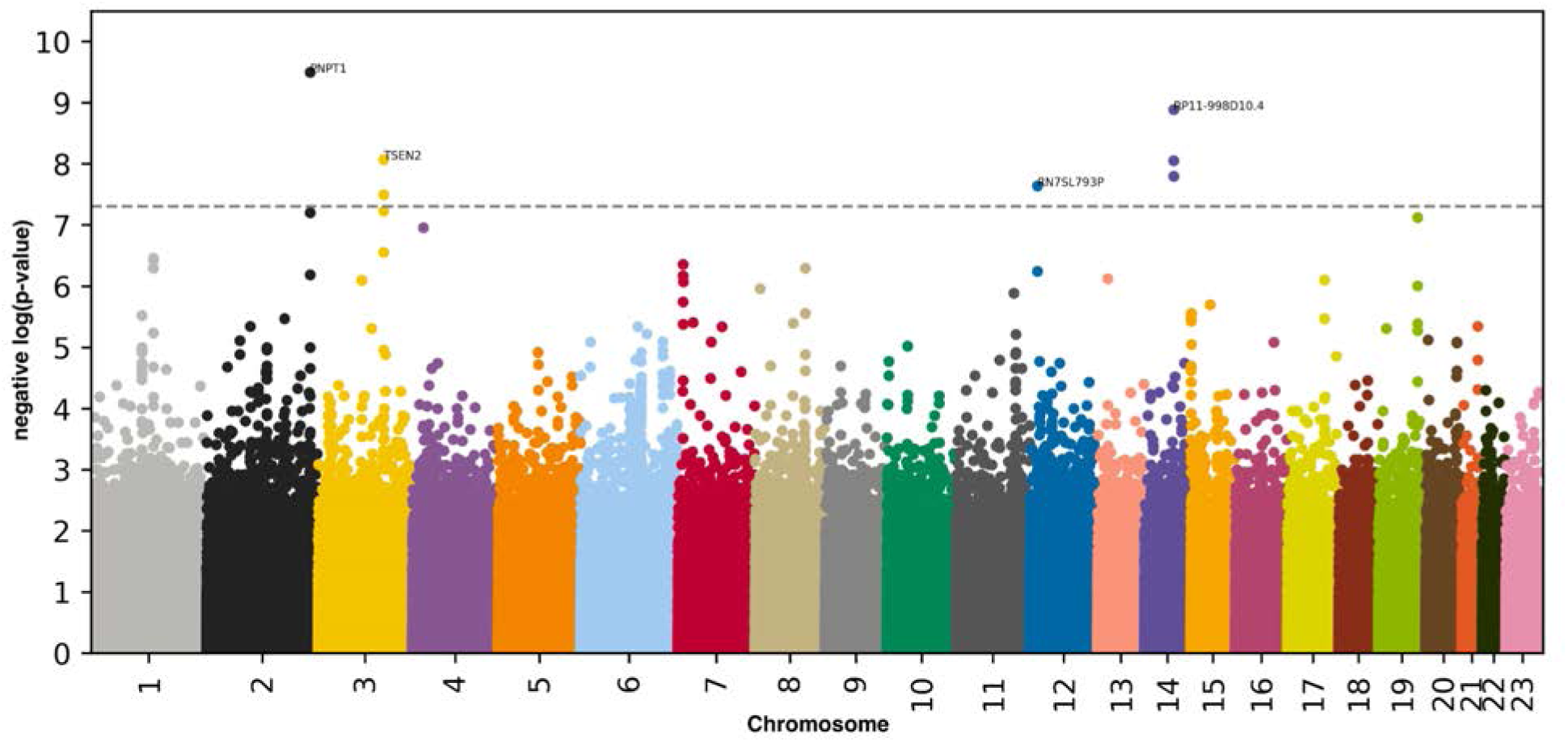
GWAS results for accelerated abdominal aging (liver MRI-based) -log10(p-value) vs. chromosomal position of locus. Dotted line denotes 5x10^-8^.

One SNP is significantly associated with general abdominal aging: PNPT1. PNPT1 is a RNA-binding protein involved in mitochondrial RNA metabolic processes. Seven SNPs in six genes are significantly associated with accelerated liver aging. The four peaks on the Manhattan plot highlight: (1) PNPT1 (Polyribonucleotide Nucleotidyltransferase 1, involved in RNA metabolic processes); (2) RP11-998D10.4 (a long intergenic non-coding RNA gene with unknown functions). Other genes in linkage disequilibrium with RP11-998D10.4 include ARHGEF40 (a protein coding gene for a guanine nucleotide exchange factor [GEF] possibly involved in G-protein-coupled receptors [GPCR] signaling) and ZNF219 (a zinc finger protein that modulates chromatin’s transcriptional activation); (3) TSEN2 (TRNA Splicing Endonuclease Subunit 2, involved in RNA splicing to remove introns); and (4) RN7SL793P (RNA, 7SL, Cytoplasmic 793, Pseudogene, a pseudogene with unknown function). ^25, 26^ We did not find any SNP significantly associated with accelerated pancreas aging.

### Biomarkers, clinical phenotypes, diseases, environmental and socioeconomic variables associated with accelerated abdominal aging

We use “X” to refer to all nongenetic variables measured in the UK Biobank (biomarkers, clinical phenotypes, diseases, family history, environmental and socioeconomic variables). We performed an X-Wide Association Study [XWAS] to identify which of the 4,372 biomarkers classified in 21 subcategories (Table S2), 187 clinical phenotypes classified in 11 subcategories (Table S5), 2,073 diseases classified in 26 subcategories (Table S8), 92 family history variables (Table S11), 265 environmental variables classified in nine categories (Table S14), and 91 socioeconomic variables classified in five categories (Table S17) are associated (p-value threshold of 0.05 and Bonferroni correction) with accelerated abdominal aging in the different dimensions. We summarize our findings for general accelerated abdominal aging below. Please refer to the supplementary tables (Table S3, Table S4, Table S6, Table S7, Table S9, Table S10, Table S15, Table S16, Table S18, Table S19) for a summary of non-genetic factors associated with general, liver MRI-based and pancreas MRI-based accelerated abdominal aging. The full results can be exhaustively explored at https://www.multidimensionality-of-aging.net/xwas/univariate_associations.

#### Biomarkers associated with accelerated abdominal aging

The three biomarker categories most associated with accelerated abdominal aging are body impedance, blood pressure, and pulse wave analysis. Specifically, 100.0% of impedance biomarkers are associated with accelerated abdominal aging, with the three largest associations being with right arm impedance (correlation=.056), left arm impedance (correlation=.055), and whole body impedance (correlation=.042). 66.7% of blood pressure biomarkers are associated with accelerated abdominal aging, with the two associations being with diastolic blood pressure (correlation=.050) and systolic blood pressure (correlation=.036). 46.7% of pulse wave analysis biomarkers are associated with accelerated abdominal aging, with the three largest associations being with diastolic blood pressure (correlation=.050), systolic blood pressure (correlation=.048), and mean arterial pressure (correlation=.046).

Conversely, the three biomarker categories most associated with decelerated abdominal aging are hand grip strength, cognitive symbol digit substitution and bone heel densitometry. Specifically, 100.%% of hand grip strength biomarkers are associated with decelerated abdominal aging, with the two associations being with left and right hand grip strengths (respective correlations of .056 and .049). 100.0% of symbol digit substitution (a cognitive test) biomarkers are associated with decelerated abdominal aging, with the two associations being with the number of symbol digit matches made correctly (correlation=.036) and the number of symbol digit matches attempted (correlation=.035). 83.3% of heel bone densitometry biomarkers are associated with decelerated abdominal aging, with the three largest associations being with heel quantitative ultrasound index (correlation=.091), heel bone mineral density (correlation=.090), and speed of sound through heel (correlation=.089).

#### Clinical phenotypes associated with accelerated abdominal aging

The three clinical phenotype categories most associated with accelerated abdominal aging are general health, chest pain and breathing. Specifically, 50.0% of general health phenotypes are associated with accelerated abdominal aging, with the three largest associations being with overall health rating (correlation=.069), weight loss in the last year (correlation=.065), and long-standing illness, disability or infirmity (correlation=.050). 50.0% of chest pain phenotypes are associated with accelerated abdominal aging, with the two associations being with chest pain or discomfort walking normally (correlation=.032) and chest pain due to walking ceasing when standing still (correlation=.023). 50.0% of breathing phenotypes are associated with accelerated abdominal aging (one association: shortness of breath walking on level ground; correlation=.031).

Conversely, the two clinical phenotype categories associated with decelerated abdominal aging are sexual factors (age first had sexual intercourse; correlation=.030) and general health (gained weight or no weight change in the last year, respective correlations of .032 and .024).

#### Diseases associated with accelerated abdominal aging

The three disease categories most associated with accelerated abdominal aging are cardiovascular diseases, general health and pulmonary diseases. Specifically, 6.5% of cardiovascular diseases are associated with accelerated abdominal aging, with the three largest associations being with hypertension (correlation=.058), atrial fibrillation and flutter (correlation=.045), and chronic ischaemic heart disease (correlation=.029). 6.0% of general health variables are associated with accelerated abdominal aging, with the three largest associations being with personal history of disease (correlation=.046), personal history of medical treatment (correlation=.042), and receiving medical care (correlation=.030). 4.8% of pulmonary diseases are associated with accelerated abdominal aging, with the three largest associations being with chronic obstructive pulmonary disease (correlation=.034), asthma (correlation=.026), and pleural effusion (correlation=.024).

We found no disease associated with decelerated abdominal aging.

#### Environmental variables associated with accelerated abdominal aging

The three environmental variable categories most associated with accelerated abdominal aging are smoking, sun exposure and alcohol intake. Specifically, 37.5% of smoking variables are associated with accelerated abdominal aging, with the three largest associations being with pack years adult smoking as proportion of lifespan exposed to smoking (correlation=.090), pack years of smoking (correlation=.086), and past tobacco smoking: smoked on most or all days (correlation=.066). 20.0% of sun exposure variables are associated with accelerated abdominal aging, with the three largest associations being with facial aging: about your age (correlation=.039), facial aging: do not know (correlation=.038), and time spent outdoors in summer (correlation=.036). 17.2% of alcohol intake variables are associated with accelerated abdominal aging, with the three largest associations being with red wine intake (correlation=.043), champagne plus white wine intake (correlation=.043), and beer plus cider intake (correlation=.042).

Conversely, the three environmental variable categories most associated with decelerated abdominal aging are physical activity, smoking and diet. Specifically, 34.3% of physical activity variables are associated with decelerated abdominal aging, with the three largest associations being with practicing strenuous sports (correlation=.078), frequency of strenuous sports in the last four weeks (correlation=.077), and duration of strenuous sports (correlation=.076). 29.2% of smoking variables are associated with decelerated abdominal aging, with the three largest associations being with smoking status: never (correlation=.073), time from waking to first cigarette (correlation=.063), and age started smoking (correlation=.062). 7.0% of diet variables are associated with decelerated abdominal aging, with the three largest associations being with cereal intake (correlation=.058), no major dietary changes in the five years (correlation=.036), and bread intake (correlation=.030).

#### Socioeconomic variables associated with accelerated abdominal aging

The two socioeconomic variable categories that are associated with accelerated abdominal aging are social support (no leisure or social activity among the ones listed: correlation=.033) and household (renting from local authority, local council or housing association: correlation=.028).

Conversely, the three socioeconomic variable categories most associated with decelerated abdominal aging are sociodemographics, employment and education. Specifically, 14.3% of sociodemographics variables are associated with decelerated abdominal aging (one association: not receiving attendance/disability/mobility allowance. correlation=.040). 13.0% of employment variables are associated with decelerated abdominal aging, with the three largest associations being with length of working week for main job (correlation=.044), current employment status: in paid employment or self-employed (correlation=.043), and frequency of travelling from home to job workplace (correlation=.029). 12.5% of education variables are associated with decelerated abdominal aging (one association: college or university degree; correlation=.048).

### Predicting accelerated abdominal aging from biomarkers, clinical phenotypes, diseases, environmental variables and socioeconomic variables

We predicted accelerated abdominal aging using variables from the different X-datasets categories (biomarkers, clinical phenotypes, diseases, environmental variables and socioeconomic variables). Specifically we built a model using the variables from each of their respective subcategories (e.g blood pressure biomarkers), and found that no dataset could explain more than 5% of the variance in accelerated abdominal aging.

### Phenotypic, genetic, and environmental correlation between liver MRI-based and pancreas MRI-based accelerated abdominal aging

Liver MRI-based and pancreas MRI-based accelerated abdominal aging are phenotypically .526±.005 correlated. For comparison purposes, the ensembles models trained on two datasets that differ only in their preprocessing (raw vs. contrasted images) yielded accelerated abdominal aging definitions that are .810±.001 correlated (liver MRIs) and .841±.002 correlated (pancreas MRIs). Liver MRI-based and pancreas MRI-based accelerated abdominal aging are genetically .863±.036 correlated. We also evaluated the correlation between liver MRI-based and pancreas MRI-based accelerated aging phenotypes in terms of their association with non-genetic variables. For example, are the environmental exposures associated with liver MRI-based accelerated aging the same as the ones associated with pancreas MRI-based accelerated aging? We found that the correlation between these two phenotypes to be .959 in terms of biomarkers, .926 in terms of associated clinical phenotypes, .793 in terms of diseases, .978 in terms of environmental variables and .969 in terms of socioeconomic variables (Figure 6). These results can be interactively explored at https://www.multidimensionality-of-aging.net/correlation_between_aging_dimensions/xwas_univariate.

**Figure 6:**
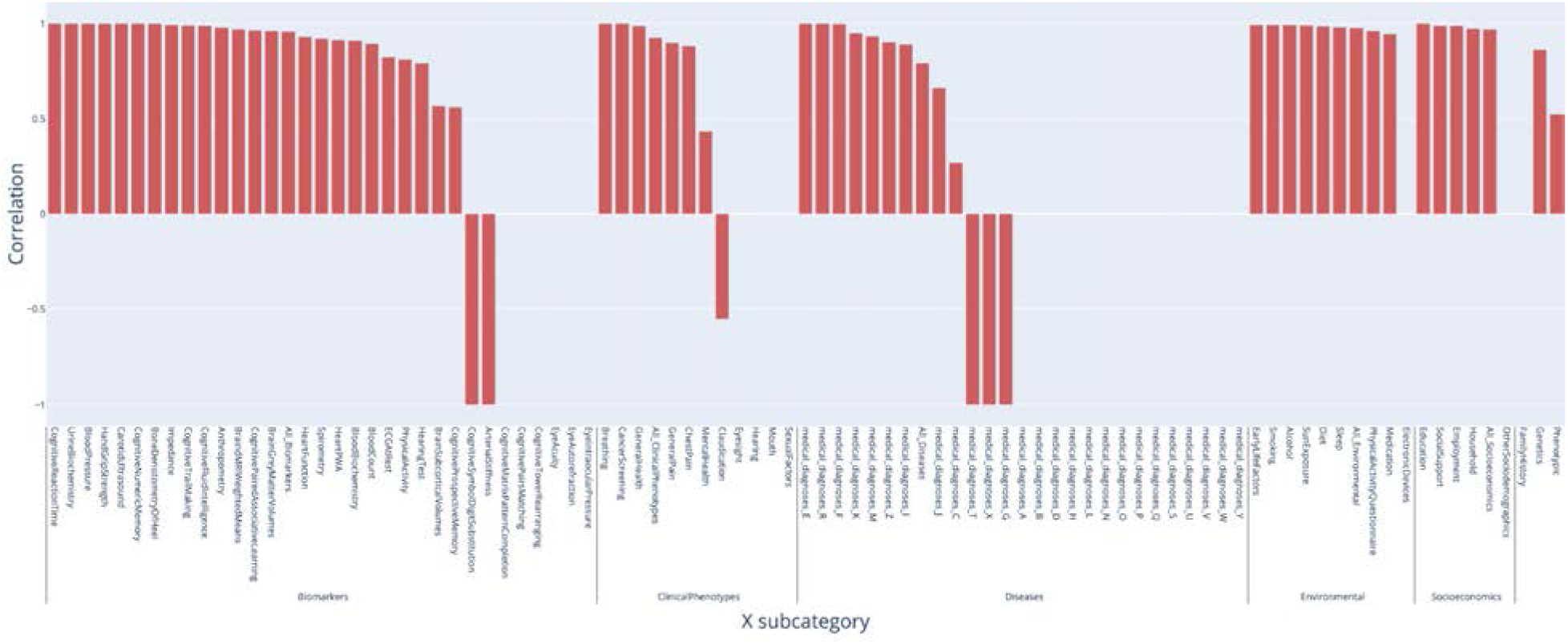
Correlation between liver MRI-based and pancreas MRI-based accelerated abdominal aging in terms of associated biomarkers, clinical phenotypes, diseases, family history, environmental and socioeconomic variables.

## Discussion

We built the first abdominal age predictor by training deep convolutional neural networks to predict age from liver and pancreas MRI images (R^2^=73.3±0.6; RMSE=3.70±0.03).

The attention maps of the models built on liver MRI images highlighted various abdominal regions including the liver, the stomach, the spleen, as well as muscle, bones and adipose tissue. The attention maps of the models built on pancreas MRI images highlighted similar features aside from the pancreas, including the liver. The similarities between liver-based and pancreas-based attention maps suggest that our models do not properly capture liver aging and pancreas aging, but instead capture general abdominal aging. The abdomen undergoes significant macroscopic changes as we age ^1^, which were likely leveraged by the convolutional neural networks. In terms of liver aging, it is known that liver function decreases with age ^27^ and that the liver ages at the cellular level, which is for example associated with low-grade inflammation ^28^. There is less evidence that the liver undergoes clear macroscopic changes that could be captured by MRI images ^29^, but it has been reported that with age the color of the liver gets darker, blood flow decreases, liver volume decreases ^3, 30^, and that the prevalence of liver diseases, such as nonalcoholic fatty liver disease, alcoholic liver disease, cirrhosis and fibrosis, increase with age ^2^, which might have been leveraged by our models to predict chronological age ^31^. In terms of pancreas aging, age-related changes visible on MRI images include pancreatic atrophy, fatty degeneration, and lobulation ^32^. Finally, aging is also associated with abdominal changes in adipose tissue ^33, 34^, muscles ^35–37^ and bones ^38^.

Further confirming the intuition derived from the attention maps, liver MRI-based and pancreas-MRI based accelerated aging are phenotypically, genetically and environmentally correlated (respective correlations of .526, .863 and .978). As a consequence, the liver MRI-based age abdominal predictor should not be interpreted as a liver age predictor. Neither should the pancreas MRI-based abdominal age predictor be considered a pancreas age predictor. To build such organ-specific predictors, it would be necessary to perform image segmentation to isolate the liver and pancreas features from their surrounding tissues and organs. Despite this limitation, liver and pancreas images captured non-redundant information regarding abdominal aging, as demonstrated by (1) the gain of prediction accuracy when combining both models (R^2^=73.3±0.6 vs. 71.5±0.6%) and (2) by the differences in GWAS signals. Specifically, the ensemble model highlighted *PNPT1* as associated with general abdominal aging, but this association was not found for pancreas MRI-based accelerated aging, despite similar sample sizes for the analysis (32,475 vs. 32,548). This difference, along with the fact that *PNPT1* was also associated with liver MRI-based accelerated aging, strongly suggests that this association is driven by features observable on liver MRIs and not on pancreas MRI.

We found that accelerated abdominal aging is heritable (h_g^2^=26.3±1.9%) and associated with seven SNPs in six different genes. The strongest association is with the *PNPT1* gene, linked to mitochondria RNA metabolic function and associated with diseases such as mitochondrial disorders, hearing loss ^39^ and Leigh syndrome ^25, 40^. The locus itself (rs10199082) has been associated with pulse pressure ^41^. We found a weaker association of loci within the *TSEN2* gene, which has been identified in GWASs in phenotypes such as body mass index adjusted waist-to-hip ratio ^42^ and may implicate a role of central adiposity of the predictor.

The association between decelerated abdominal aging and blood biochemistry biomarkers such as alanine aminotransferase, aspartate aminotransferase suggest that abdominal aging is linked to liver function. Since age prediction is in part driven by the tissue surrounding the organs, a natural hypothesis is that the model also relies on body/liver fat percentage, which is known to increase with age ^43^. This hypothesis is partly confirmed by the fact that the biomarker category most associated with accelerated aging is body impedance, which increases with body fat percentage. Similarly, metabolism biomarkers such as HDL cholesterol, apolipoprotein A and glycated haemoglobin A1c (a diabetes biomarker) are associated with accelerated abdominal aging. However, and perhaps surprisingly, both body mass index, hip circumference and weight are associated with decelerated abdominal aging. A possible explanation is that both old age ^44^ and disease (e.g pancreas cancer ^45^, cirrhosis ^46^) are associated with weight loss.

Aside from these biomarkers which can be linked to abdominal health, accelerated abdominal aging is also associated with biomarkers, clinical phenotypes and diseases linked to other organ systems’ health that cannot be not directly observed from liver and pancreas MRIs. For example, it is associated with poor cardiovascular health (e.g blood pressure, chest pain, hypertension, atrial fibrillation and flutter, chronic ischaemic heart disease), brain health (cognitive tests, brain MRI volumes, mental health disorders such as fed up feelings and mood swings), and pulmonary function (e.g spirometry, shortness of breath, chronic obstructive pulmonary disease, asthma and pleural effusion). More generally, accelerated abdominal aging is associated with poor general health (e.g general health rating, recent weight loss, long-standing illness, disability or infirmity, personal history of disease and medical treatment), suggesting that accelerated aging in the different organ systems is linked. We explore this hypothesis of the multidimensionality of aging in a different paper ^47^. Interestingly, accelerated abdominal aging is also correlated with facial aging.

In terms of environmental variables, we found that smoking and sedentarity (e.g time spent watching television, lack of strenuous physical activity) are associated with accelerated abdominal aging, in accordance with the unambiguous literature on the subject ^48, 49^. We found some diet variables to be associated with decelerated abdominal aging (e.g cereal intake, bread intake). More generally, having a stable weight was associated with decelerated aging. Alcohol had a mixed association, with champagne, white wine, beer, cider and red wine intake being all associated with accelerated abdominal aging, while alcohol intake frequency was associated with decelerated abdominal aging, possibly reflecting the complex literature on the topic ^50^. Socioeconomic status (e.g education, income) was also negatively correlated with accelerated abdominal aging. In a developed country such as the US, the richest 1% live more than a decade longer than their poorest 1% counterparts, on average (10.1±0.2 years for females, 14.6±0.2 years longer for males) ^51^. This difference could be mediated by better access to healthcare and health literacy ^52^.

In conclusion, our biological age predictor can be used to assess abdominal aging and defines an accelerated aging phenotype that is linked to disease. Our GWAS and XWAS findings suggest potential lifestyle and therapeutic interventions to slow or reverse abdominal aging. Additionally, our predictor could be used on clinical trials to assess the effect of emerging rejuvenating therapies ^53^ on abdominal organs and tissue. Other age predictors such as the DNA methylation clock are already leveraged to this end ^18, 54, 55^ but, as aging is multidimensional ^47, 56^, diverse predictors will be needed to fully measure the therapeutic effect of candidate drugs on the different organs and tissues.

## Methods

### Data and materials availability

We used the UK Biobank (project ID: 52887). The code can be found at https://github.com/Deep-Learning-and-Aging. The results can be interactively and extensively explored at https://www.multidimensionality-of-aging.net/. We will make the biological age phenotypes available through UK Biobank upon publication. The GWAS results can be found at https://www.dropbox.com/s/59e9ojl3wu8qie9/Multidimensionality_of_aging-GWAS_results.zip?dl=0.

### Software

Our code can be found at https://github.com/Deep-Learning-and-Aging. For the genetics analysis, we used the BOLT-LMM ^57, 58^ and BOLT-REML ^59^ softwares. We coded the parallel submission of the jobs in Bash ^60^.

### Cohort Dataset: Participants of the UK Biobank

We leveraged the UK Biobank^24^ cohort (project ID: 52887). The UKB cohort consists of data originating from a large biobank collected from 502,211 de-identified participants in the United Kingdom that were aged between 37 years and 74 years at enrollment (starting in 2006). Out of these participants, 44,481 had liver MRIs collected from them, and 36,591 had pancreas MRIs collected from them. The Harvard internal review board (IRB) deemed the research as non-human subjects research (IRB: IRB16-2145).

### Data types and Preprocessing

#### Demographic variables

First, we removed out the UKB samples for which age or sex was missing. For sex, we used the genetic sex when available, and the self-reported sex when genetic sex was not available. We computed age as the difference between the date when the participant attended the assessment center and the year and month of birth of the participant to estimate the participant’s age with greater precision. We one-hot encoded ethnicity.

#### Liver and Pancreas MRIs

UKB contains Liver MRI images (field 20204, 45,685 samples for 43,267 participants) of dimensions 288*384, stored as DICOM files. We removed the 83 images for which the image quality indicator had any flag on (field 22414). We applied an adaptive histogram equalizer filter to the images to enhance the contrast. We kept both images, which we named “Raw” and “Contrast”. We cropped off the legend on the right side of the images which yielded images of dimensions 288*350, that we stored as .jpg images. The UKB also contains pancreas images (field 20259, 37,619 samples for 35,285 participants). We followed the same pipeline used for the preprocessing of the liver images to preprocess the pancreas images and obtained 36,784 images. A sample of preprocessed abdominal (liver and pancreas) images can be found in Figure 1A.

#### Data augmentation

To prevent overfitting and increase our sample size during the training we used data augmentation ^61^ on the images. Each image was randomly shifted vertically (maximal amplitude ±10%) and horizontally (maximal amplitude ±10%), as well as rotated (maximal angle ± 10 degrees). We chose the hyperparameters for these transformations’ distributions to represent the variations we observed between the images in the initial dataset. For example, we observed similar variation between images in the vertical and the horizontal direction, so both the random vertical and horizontal shifts were sampled from the [-10%, +10%] uniform distribution.

The data augmentation process is dynamically performed during the training. Augmented images are not generated in advance. Instead, each image is randomly augmented before being fed to the neural network for each epoch during the training.

### Machine learning algorithms

#### Convolutional Neural Networks Architectures

We used transfer learning ^62–64^ to leverage two different convolutional neural networks ^65^ [CNN] architectures pre-trained on the ImageNet dataset ^66–68^ and made available through the python Keras library ^69^: InceptionV3 ^70^ and InceptionResNetV2 ^71^. We considered other architectures such as VGG16 ^72^, VGG19 ^72^ and EfficientNetB7 ^73^, but found that they performed poorly and inconsistently on our datasets during our preliminary analysis and we therefore did not train them in the final pipeline. For each architecture, we removed the top layers initially used to predict the 1,000 different ImageNet images categories. We refer to this truncated model as the “base CNN architecture”.

We added to the base CNN architecture what we refer to as a “side neural network”. A side neural network is a single fully connected layer of 16 nodes, taking the sex and the ethnicity variables of the participant as input. The output of this small side neural network was concatenated to the output of the base CNN architecture described above. This architecture allowed the model to consider the features extracted by the base CNN architecture in the context of the sex and ethnicity variables. For example, the presence of the same anatomical feature can be interpreted by the algorithm differently for a male and for a female. We added several sequential fully connected dense layers after the concatenation of the outputs of the CNN architecture and the side neural architecture. The number and size of these layers were set as hyperparameters. We used ReLU ^74^ as the activation function for the dense layers we added, and we regularized them with a combination of weight decay ^75, 76^ and dropout ^77^, both of which were also set as hyperparameters. Finally, we added a dense layer with a single node and linear activation to predict age.

#### Compiler

The compiler uses gradient descent ^78, 79^ to train the model. We treated the gradient descent optimizer, the initial learning rate and the batch size as hyperparameters. We used mean squared error [MSE] as the loss function, root mean squared error [RMSE] as the metric and we clipped the norm of the gradient so that it could not be higher than 1.0 ^80^.

We defined an epoch to be 32,768 images. If the training loss did not decrease for seven consecutive epochs, the learning rate was divided by two. This is theoretically redundant with the features of optimizers such as Adam, but we found that enforcing this manual decrease of the learning rate was sometimes beneficial. During training, after each image has been seen once by the model, the order of the images is shuffled. At the end of each epoch, if the validation performance improved, the model’s weights were saved.

We defined convergence as the absence of improvement on the validation loss for 15 consecutive epochs. This strategy is called early stopping ^81^ and is a form of regularization. We requested the GPUs on the supercomputer for ten hours. If a model did not converge within this time and improved its performance at least once during the ten hours period, another GPU was later requested to reiterate the training, starting from the model’s last best weights.

### Training, tuning and predictions

We split the entire dataset into ten data folds. We manually tuned some of the hyperparameters before performing a simple cross-validation. We describe the splitting of the data into different folds and the tuning procedures in greater detail in the Supplementary.

### Interpretability of the machine learning predictions

To interpret the models, we used attention maps (saliency and Grad-RAM). See supplementary Methods).

### Ensembling to improve prediction and define aging dimensions

We built a three-level hierarchy of ensemble models to improve prediction accuracies. At the lowest level, we combined the predictions from different algorithms on the same dataset. For example, we combined the predictions generated by InceptionResNetv2 and Inceptionv3 from raw liver MRI images into a single raw liver MRI-based prediction. At the second level, we combined the predictions from the different preprocessing (raw and contrasted images) into a prediction for a specific organ (liver or pancreas). For the third and highest level, we combined all predictions into a general abdomen-based prediction. The ensemble models from the lower levels are hierarchically used as components of the ensemble models of the higher models. For example, the ensemble model built by combining the algorithms trained on raw liver MRIs is leveraged when building the general abdominal aging ensemble model.

We built each ensemble model separately on each of the ten data folds. For example, to build the ensemble model on the testing predictions of the data fold #1, we trained and tuned an elastic net on the validation predictions from the data fold #0 using a 10-folds inner cross-validation, as the validation predictions on fold #0 and the testing predictions on fold #1 are generated by the same model. We used the same hyperparameters space and Bayesian hyperparameters optimization method as we did for the inner cross-validation we performed during the tuning of the non-ensemble models.

To summarize, the testing ensemble predictions are computed by concatenating the testing predictions generated by ten different elastic nets, each of which was trained and tuned using a 10-folds inner cross-validation on one validation data fold (10% of the full dataset) and tested on one testing fold. This is different from the inner-cross validation performed when training the non-ensemble models, which was performed on the “training+validation” data folds, so on 9 data folds (90% of the dataset).

### Evaluating the performance of models

We evaluated the performance of the models using two different metrics: R-Squared [R^2^] and root mean squared error [RMSE]. We computed a confidence interval on the performance metrics in two different ways. First, we computed the standard deviation between the different data folds. The test predictions on each of the ten data folds are generated by ten different models, so this measure of standard deviation captures both model variability and the variability in prediction accuracy between samples. Second, we computed the standard deviation by bootstrapping the computation of the performance metrics 1,000 times. This second measure of variation does not capture model variability but evaluates the variance in the prediction accuracy between samples.

### Abdominal age definition

We defined the abdominal age of participants for a specific abdominal dimension as the prediction outputted by the model trained on the corresponding dataset, after correcting for the bias in the residuals.

We indeed observed a bias in the residuals. For each model, participants on the older end of the chronological age distribution tend to be predicted younger than they are. Symmetrically, participants on the younger end of the chronological age distribution tend to be predicted older than they are. This bias does not seem to be biologically driven. Rather it seems to be statistically driven, as the same 60-year-old individual will tend to be predicted younger in a cohort with an age range of 60-80 years, and to be predicted older in a cohort with an age range of 60-80. We ran a linear regression on the residuals as a function of age for each model and used it to correct each prediction for this statistical bias.

After defining biological age as the corrected prediction, we defined accelerated aging as the corrected residuals. For example, a 60-year-old whose liver MRI predicted an age of 70 years old after correction for the bias in the residuals is estimated to have a liver MRI-based abdominal age of 70 years, and an accelerated abdominal aging of ten years.

It is important to understand that this step of correction of the predictions and the residuals takes place after the evaluation of the performance of the models but precedes the analysis of the abdominal ages properties.

### Genome-wide association of accelerated abdominal aging

The UKB contains genome-wide genetic data for 488,251 of the 502,492 participants^82^ under the hg19/GRCh37 build.

We used the average accelerated aging value over the different samples collected over time (see Supplementary - Generating average predictions for each participant). Next, we performed genome wide association studies [GWASs] to identify single-nucleotide polymorphisms [SNPs] associated with accelerated aging in each abdominal dimension using BOLT-LMM ^57, 58^ and estimated the the SNP-based heritability for each of our biological age phenotypes, and we computed the genetic pairwise correlations between dimensions using BOLT-REML ^59^. We used the v3 imputed genetic data to increase the power of the GWAS, and we corrected all of them for the following covariates: age, sex, ethnicity, the assessment center that the participant attended when their DNA was collected, and the 20 genetic principal components precomputed by the UKB. We used the linkage disequilibrium [LD] scores from the 1,000 Human Genomes Project ^83^. To avoid population stratification, we performed our GWAS on individuals with White ethnicity.

#### Identification of SNPs associated with accelerated abdominal aging

We identified the SNPs associated with accelerated abdominal aging dimensions using the BOLT-LMM ^57, 58^ software (p-value of 5e-8). The sample size for the genotyping of the X chromosome is one thousand samples smaller than for the autosomal chromosomes. We therefore performed two GWASs for each aging dimension. (1) excluding the X chromosome, to leverage the full autosomal sample size when identifying the SNPs on the autosome. (2) including the X chromosome, to identify the SNPs on this sex chromosome. We then concatenated the results from the two GWASs to cover the entire genome, at the exception of the Y chromosome.

We plotted the results using a Manhattan plot and a volcano plot. We used the bioinfokit ^84^ python package to generate the Manhattan plots. We generated quantile-quantile plots [Q-Q plots] to estimate the p-value inflation as well.

#### Heritability and genetic correlation

We estimated the heritability of the accelerated aging dimensions using the BOLT-REML ^59^ software. We included the X chromosome in the analysis and corrected for the same covariates as we did for the GWAS. Using the same software and parameters, we computed the genetic correlations between accelerated aging in the two image-based abdominal dimensions.

We annotated the significant SNPs with their matching genes using the following four steps pipeline. (1) We annotated the SNPs based on the rs number using SNPnexus ^85–89^. When the SNP was between two genes, we annotated it with the nearest gene. (2) We used SNPnexus to annotate the SNPs that did not match during the first step, this time using their genomic coordinates. After these two first steps, 30 out of the 9,697 significant SNPs did not find a match. (3) We annotated these SNPs using LocusZoom ^90^. Unlike SNPnexus, LocusZoom does not provide the gene types, so we filled this information with GeneCards ^26^. After this third step, four genes were not matched. (4) We used RCSB Protein Data Bank ^91^ to annotate three of the four missing genes.

### Non-genetic correlates of accelerated abdominal aging

We identified non-genetically measured (i.e factors not measured on a GWAS array) correlates of each aging dimension, which we classified in six categories: biomarkers, clinical phenotypes, diseases, family history, environmental, and socioeconomic variables. We refer to the union of these association analyses as an X-Wide Association Study [XWAS]. (1) We define as biomarkers the scalar variables measured on the participant, which we initially leveraged to predict age (e.g. blood pressure, Table S2). (2) We define clinical phenotypes as other biological factors not directly measured on the participant, but instead collected by the questionnaire, and which we did not use to predict chronological age. For example, one of the clinical phenotypes categories is eyesight, which contains variables such as “wears glasses or contact lenses”, which is different from the direct refractive error measurements performed on the participants, which are considered “biomarkers” (Table S5). (3) Diseases include the different medical diagnoses categories listed by UKB (Table S8). (4) Family history variables include illnesses of family members (Table S11). (5) Environmental variables include alcohol, diet, electronic devices, medication, sun exposure, early life factors, medication, sun exposure, sleep, smoking, and physical activity variables collected from the questionnaire (Table S14). (6) Socioeconomic variables include education, employment, household, social support and other sociodemographics (Table S17). We provide information about the preprocessing of the XWAS in the Supplementary Methods.

## Data Availability

https://www.multidimensionality-of-aging.net/

https://www.dropbox.com/s/59e9ojl3wu8qie9/Multidimensionality_of_aging-GWAS_results.zip?dl=0

## Author Contributions

**Alan Le Goallec:** (1) Designed the project. (2) Supervised the project. (3) Predicted chronological age from liver and pancreas MRIs. (4) Computed the attention maps for the images. (5) Ensembled the models, evaluated their performance, computed biological ages and estimated the correlation structure between the abdominal aging dimensions. (6) Performed the genome wide association studies. (5) Designed the website. (6) Wrote the manuscript.

**Samuel Diai:** (1) Wrote the python class to build an ensemble model using a cross-validated elastic net. (2) Performed the X-wide association study. (3) Implemented a first version of the website https://www.multidimensionality-of-aging.net/.

**Jean Baptiste Prost:** (1) Preprocessed the liver images.

**Sasha Collin:** (1) Preprocessed the pancreas images.

**Théo Vincent:** (1) Website data engineer. (2) Implemented a second version of the website https://www.multidimensionality-of-aging.net/.

**Chirag J. Patel:** (1) Supervised the project. (2) Edited the manuscript. (3) Provided funding.

## Acknowledgments

We would like to thank Raffaele Potami from Harvard Medical School research computing group for helping us utilize O2’s computing resources. We thank HMS RC for computing support. We also want to acknowledge UK Biobank for providing us with access to the data they collected. The UK Biobank project number is 52887.

## Conflicts of Interest

None.

## Funding

NIEHS R00 ES023504

NIEHS R21 ES25052.

NIAID R01 AI127250

NSF 163870

MassCATS, Massachusetts Life Science Center Sanofi

The funders had no role in the study design or drafting of the manuscript(s).

## Notes

### Competing Interest Statement

The authors have declared no competing interest.

### Author Declarations

The Harvard internal review board (IRB) deemed the research as non-human subjects research (IRB: IRB16-2145).

## References

1. Meier, J. M. et al. Assessment of age-related changes in abdominal organ structure and function with computed tomography and positron emission tomography. Semin. Nucl. Med. 37, 154–172 (2007).

2. Kim, I. H., Kisseleva, T. & Brenner, D. A. Aging and liver disease. Curr. Opin. Gastroenterol. 31, 184–191 (2015).

3. Schmucker, D. L. Age-related changes in liver structure and function: Implications for disease ? Exp. Gerontol. 40, 650–659 (2005).

4. Matsuda, Y. Age-related pathological changes in the pancreas. Front. Biosci. 10, 137–142 (2018).

5. Matsuda, Y. Age-related morphological changes in the pancreas and their association with pancreatic carcinogenesis. Pathol. Int. 69, 450–462 (2019).

6. Löhr, J.-M., Panic, N., Vujasinovic, M. & Verbeke, C. S. The ageing pancreas: a systematic review of the evidence and analysis of the consequences. J. Intern. Med. 283, 446–460 (2018).

7. Soenen, S., Rayner, C. K., Jones, K. L. & Horowitz, M. The ageing gastrointestinal tract. Curr. Opin. Clin. Nutr. Metab. Care 19, 12–18 (2016).

8. Dinsdale, N. K. et al. Learning patterns of the ageing brain in MRI using deep convolutional networks. Neuroimage 224, 117401 (2021).

9. Le Goallec, A. et al. Dissecting heart age using cardiac magnetic resonance videos, electrocardiograms, biobanks, and deep learning. medRxiv (2021) doi:10.1101/2021.06.09.21258645.

10. Attia, Z. I. et al. Age and Sex Estimation Using Artificial Intelligence From Standard 12-Lead ECGs. Circ. Arrhythm. Electrophysiol. 12, e007284 (2019).

11. Goallec, A. L. et al. Predicting arterial age using carotid ultrasound images, pulse wave analysis records, cardiovascular biomarkers and deep learning. doi:10.1101/2021.06.17.21259120.

12. Goallec, A. L. et al. Using deep learning to analyze the compositeness of musculoskeletal aging reveals that spine, hip and knee age at different rates, and are associated with different genetic and non-genetic factors. doi:10.1101/2021.06.14.21258896.

13. Langner, T., Wikstrom, J., Bjerner, T., Ahlstrom, H. & Kullberg, J. Identifying Morphological Indicators of Aging With Neural Networks on Large-Scale Whole-Body MRI. IEEE Trans. Med. Imaging 39, 1430–1437 (2020).

14. Karargyris, A. et al. Age prediction using a large chest x-ray dataset. Medical Imaging 2019: Computer-Aided Diagnosis (2019) doi:10.1117/12.2512922.

15. Poplin, R. et al. Prediction of cardiovascular risk factors from retinal fundus photographs via deep learning. Nat Biomed Eng 2, 158–164 (2018).

16. Smith, P. & Chen, C. Transfer Learning with Deep CNNs for Gender Recognition and Age Estimation. 2018 IEEE International Conference on Big Data (Big Data) (2018) doi:10.1109/bigdata.2018.8621891.

17. Putin, E. et al. Deep biomarkers of human aging: Application of deep neural networks to biomarker development. Aging 8, 1021–1033 (2016).

18. Horvath, S. DNA methylation age of human tissues and cell types. Genome Biol. 14, R115 (2013).

19. Mamoshina, P. et al. Machine Learning on Human Muscle Transcriptomic Data for Biomarker Discovery and Tissue-Specific Drug Target Identification. Frontiers in Genetics vol. 9 (2018).

20. Lehallier, B., Shokhirev, M. N., Wyss-Coray, T. & Johnson, A. A. Data mining of human plasma proteins generates a multitude of highly predictive aging clocks that reflect different aspects of aging. Aging Cell e13256 (2020).

21. Galkin, F. et al. Human Gut Microbiome Aging Clock Based on Taxonomic Profiling and Deep Learning. iScience 23, 101199 (2020).

22. Le Goallec, A. et al. A systematic machine learning and data type comparison yields metagenomic predictors of infant age, sex, breastfeeding, antibiotic usage, country of origin, and delivery type. PLoS Comput. Biol. 16, e1007895 (2020).

23. Rahman, S. A. & Adjeroh, D. A. Deep Learning using Convolutional LSTM estimates Biological Age from Physical Activity. Sci. Rep. 9, 11425 (2019).

24. Sudlow, C. et al. UK biobank: an open access resource for identifying the causes of a wide range of complex diseases of middle and old age. PLoS Med. 12, e1001779 (2015).

25. Safran, M. et al. GeneCards Version 3: the human gene integrator. Database 2010, baq020 (2010).

26. Stelzer, G. et al. The GeneCards Suite: From Gene Data Mining to Disease Genome Sequence Analyses. Curr. Protoc. Bioinformatics 54, 1.30.1–1.30.33 (2016).

27. Cieslak, K. P., Baur, O., Verheij, J., Bennink, R. J. & van Gulik, T. M. Liver function declines with increased age. HPB 18, 691–696 (2016).

28. Hunt, N. J., Kang, S. W. S., Lockwood, G. P., Le Couteur, D. G. & Cogger, V. C. Hallmarks of Aging in the Liver. Comput. Struct. Biotechnol. J. 17, 1151–1161 (2019).

29. Pasquinelli, F., Belli, G., Mazzoni, L. N., Grazioli, L. & Colagrande, S. Magnetic resonance diffusion-weighted imaging: quantitative evaluation of age-related changes in healthy liver parenchyma. Magn. Reson. Imaging 29, 805–812 (2011).

30. Woodhouse, K. W. & Wynne, H. A. Age-related changes in liver size and hepatic blood flow. The influence on drug metabolism in the elderly. Clin. Pharmacokinet. 15, 287–294 (1988).

31. Chundru, S. et al. MRI of diffuse liver disease: characteristics of acute and chronic diseases. Diagn. Interv. Radiol. 20, 200–208 (2014).

32. Sato, T. et al. Age-related changes in normal adult pancreas: MR imaging evaluation. European Journal of Radiology vol. 81 2093–2098 (2012).

33. Mancuso, P. & Bouchard, B. The Impact of Aging on Adipose Function and Adipokine Synthesis. Front. Endocrinol. 10, 137 (2019).

34. Hunter, G. R. et al. Weight loss needed to maintain visceral adipose tissue during aging. Int. J. Body Compos. Res. 3, 55 (2005).

35. Ota, M., Ikezoe, T., Kato, T., Tateuchi, H. & Ichihashi, N. Age-related changes in muscle thickness and echo intensity of trunk muscles in healthy women: comparison of 20–60s age groups. European Journal of Applied Physiology vol. 120 1805–1814 (2020).

36. Ota, M., Ikezoe, T., Kaneoka, K. & Ichihashi, N. Age-related changes in the thickness of the deep and superficial abdominal muscles in women. Arch. Gerontol. Geriatr. 55, e26–30 (2012).

37. Tanaka, N. I. et al. Difference in abdominal muscularity at the umbilicus level between young and middle-aged men. J. Physiol. Anthropol. 26, 527–532 (2007).

38. Benoist, M. Natural history of the aging spine. Eur. Spine J. 12 **Suppl 2**, S86–9 (2003).

39. von Ameln, S. et al. A Mutation in PNPT1, Encoding Mitochondrial-RNA-Import Protein PNPase, Causes Hereditary Hearing Loss. Am. J. Hum. Genet. 91, 919–927 (2012).

40. Matilainen, S. et al. Defective mitochondrial RNA processing due to PNPT1 variants causes Leigh syndrome. Hum. Mol. Genet. 26, 3352–3361 (2017).

41. Hoffmann, T. J. et al. Genome-wide association analyses using electronic health records identify new loci influencing blood pressure variation. Nat. Genet. 49, 54–64 (2017).

42. Lotta, L. A. et al. Association of Genetic Variants Related to Gluteofemoral vs Abdominal Fat Distribution With Type 2 Diabetes, Coronary Disease, and Cardiovascular Risk Factors. JAMA 320, 2553–2563 (2018).

43. Zamboni, M. et al. Effects of age on body fat distribution and cardiovascular risk factors in women. Am. J. Clin. Nutr. 66, 111–115 (1997).

44. Mott, J. W. et al. Relation between body fat and age in 4 ethnic groups. Am. J. Clin. Nutr. 69, 1007–1013 (1999).

45. Hendifar, A. E. et al. Pancreas cancer-associated weight loss. Oncologist 24, 691–701 (2019).

46. Anastácio, L. R. et al. Weight loss during cirrhosis is related to the etiology of liver disease. Arq. Gastroenterol. 49, 195–198 (2012).

47. Le Goallec, A. et al. Analyzing the multidimensionality of biological aging with the tools of deep learning across diverse image-based and physiological indicators yields robust age predictors. medRxiv (2021).

48. Warburton, D. E. R., Nicol, C. W. & Bredin, S. S. D. Health benefits of physical activity: the evidence. CMAJ 174, 801–809 (2006).

49. Jha, P. The hazards of smoking and the benefits of cessation: a critical summation of the epidemiological evidence in high-income countries. Elife 9, (2020).

50. Burton, R. & Sheron, N. No level of alcohol consumption improves health. The Lancet vol. 392 987–988 (2018).

51. Chetty, R. et al. The Association Between Income and Life Expectancy in the United States, 2001-2014. JAMA 315, 1750–1766 (2016).

52. Liu, C. et al. What is the meaning of health literacy? A systematic review and qualitative synthesis. Family medicine and community health 8, (2020).

53. de Magalhães, J. P., Stevens, M. & Thornton, D. The Business of Anti-Aging Science. Trends Biotechnol. 35, 1062–1073 (2017).

54. Duke Clinical Research Institute, Elysium Health. Biomarker Study to Evaluate Correlations Between Epigenetic Aging and NAD+ Levels in Healthy Volunteers. (2019).

55. Horvath, S. et al. Obesity accelerates epigenetic aging of human liver. Proc. Natl. Acad. Sci. U. S. A. 111, 15538–15543 (2014).

56. Li, X. et al. Longitudinal trajectories, correlations and mortality associations of nine biological ages across 20-years follow-up. eLife vol. 9 (2020).

57. Loh, P.-R. et al. Efficient Bayesian mixed-model analysis increases association power in large cohorts. Nat. Genet. 47, 284–290 (2015).

58. Loh, P.-R., Kichaev, G., Gazal, S., Schoech, A. P. & Price, A. L. Mixed-model association for biobank-scale datasets. Nat. Genet. 50, 906–908 (2018).

59. Loh, P.-R. et al. Contrasting genetic architectures of schizophrenia and other complex diseases using fast variance-components analysis. Nature Genetics vol. 47 1385–1392 (2015).

60. Gnu, P. Free Software Foundation. Bash (3. 2. 48)[Unix shell program] (2007).

61. Shorten, C. & Khoshgoftaar, T. M. A survey on Image Data Augmentation for Deep Learning. Journal of Big Data 6, 60 (2019).

62. Tan, C. et al. A Survey on Deep Transfer Learning. in Artificial Neural Networks and Machine Learning – ICANN 2018 270–279 (Springer International Publishing, 2018).

63. Weiss, K., Khoshgoftaar, T. M. & Wang, D. A survey of transfer learning. Journal of Big data 3, 9 (2016).

64. Pan, S. J. & Yang, Q. A Survey on Transfer Learning. IEEE Trans. Knowl. Data Eng. 22, 1345–1359 (2010).

65. LeCun, Y., Bengio, Y. & Hinton, G. Deep learning. Nature vol. 521 436–444 (2015).

66. Deng, J. et al. ImageNet: A large-scale hierarchical image database. in 2009 IEEE Conference on Computer Vision and Pattern Recognition 248–255 (2009).

67. Krizhevsky, A., Sutskever, I. & Hinton, G. E. ImageNet Classification with Deep Convolutional Neural Networks. in Advances in Neural Information Processing Systems 25 (eds. Pereira, F., Burges, C. J. C., Bottou, L. & Weinberger, K. Q.) 1097–1105 (Curran Associates, Inc., 2012).

68. Russakovsky, O. et al. ImageNet Large Scale Visual Recognition Challenge. Int. J. Comput. Vis. 115, 211–252 (2015).

69. Chollet, F. & Others. keras. (2015).

70. Szegedy, C., Vanhoucke, V., Ioffe, S., Shlens, J. & Wojna, Z. Rethinking the inception architecture for computer vision. in Proceedings of the IEEE conference on computer vision and pattern recognition 2818–2826 (2016).

71. Szegedy, C., Ioffe, S., Vanhoucke, V. & Alemi, A. A. Inception-v4, inception-resnet and the impact of residual connections on learning. in Thirty-first AAAI conference on artificial intelligence (2017).

72. Simonyan, K. & Zisserman, A. Very Deep Convolutional Networks for Large-Scale Image Recognition. arXiv [cs.CV] (2014).

73. Tan, M. & Le, Q. V. EfficientNet: Rethinking Model Scaling for Convolutional Neural Networks. arXiv [cs.LG] (2019).

74. Agarap, A. F. Deep Learning using Rectified Linear Units (ReLU). arXiv [cs.NE] (2018).

75. Krogh, A. & Hertz, J. A. A Simple Weight Decay Can Improve Generalization. in Advances in Neural Information Processing Systems 4 (eds. Moody, J. E., Hanson, S. J. & Lippmann, R. P.) 950–957 (Morgan-Kaufmann, 1992).

76. Bos, S. & Chug, E. Using weight decay to optimize the generalization ability of a perceptron. Proceedings of International Conference on Neural Networks (ICNN’96) doi:10.1109/icnn.1996.548898.

77. Srivastava, N., Hinton, G., Krizhevsky, A., Sutskever, I. & Salakhutdinov, R. Dropout: a simple way to prevent neural networks from overfitting. J. Mach. Learn. Res. 15, 1929–1958 (2014).

78. Ruder, S. An overview of gradient descent optimization algorithms. arXiv [cs.LG] (2016).

79. Bottou, L., Curtis, F. E. & Nocedal, J. Optimization Methods for Large-Scale Machine Learning. SIAM Rev. 60, 223–311 (2018).

80. Zhang, J., He, T., Sra, S. & Jadbabaie, A. Why gradient clipping accelerates training: A theoretical justification for adaptivity. arXiv [math.OC] (2019).

81. Prechelt, L. Early Stopping - But When? in Neural Networks: Tricks of the Trade (eds. Orr, G. B. & Müller, K.-R.) 55–69 (Springer Berlin Heidelberg, 1998).

82. Bycroft, C. et al. Genome-wide genetic data on\ 500,000 UK Biobank participants. BioRxiv 166298 (2017).

83. Consortium, T. 1000 G. P. & The 1000 Genomes Project Consortium. A global reference for human genetic variation. Nature vol. 526 68–74 (2015).

84. Bedre, R. reneshbedre/bioinfokit: Bioinformatics data analysis and visualization toolkit. (2020). doi:10.5281/zenodo.3965241.

85. Oscanoa, J. et al. SNPnexus: a web server for functional annotation of human genome sequence variation (2020 update). Nucleic Acids Res. 48, W185–W192 (2020).

86. Dayem Ullah, A. Z. et al. SNPnexus: assessing the functional relevance of genetic variation to facilitate the promise of precision medicine. Nucleic Acids Res. 46, W109–W113 (2018).

87. Ullah, A. Z. D., Dayem Ullah, A. Z., Lemoine, N. R. & Chelala, C. A practical guide for the functional annotation of genetic variations using SNPnexus. Briefings in Bioinformatics vol. 14 437–447 (2013).

88. Dayem Ullah, A. Z., Lemoine, N. R. & Chelala, C. SNPnexus: a web server for functional annotation of novel and publicly known genetic variants (2012 update). Nucleic Acids Res. 40, W65–70 (2012).

89. Chelala, C., Khan, A. & Lemoine, N. R. SNPnexus: a web database for functional annotation of newly discovered and public domain single nucleotide polymorphisms. Bioinformatics 25, 655–661 (2009).

90. Pruim, R. J. et al. LocusZoom: regional visualization of genome-wide association scan results. Bioinformatics 26, 2336–2337 (2010).

91. Berman, H. M. et al. The protein data bank. Nucleic Acids Res. 28, 235–242 (2000).

